# Head-to-head comparison of direct-input RT-PCR and RT-LAMP against RT-qPCR on extracted RNA for rapid SARS-CoV-2 diagnostics

**DOI:** 10.1101/2021.01.19.21250079

**Authors:** Max J. Kellner, Martin Matl, James J. Ross, Jakob Schnabl, Dominik Handler, Robert Heinen, Justine Schaeffer, Peter Hufnagl, Alexander Indra, Marcus P.S. Dekens, Robert Fritsche-Polanz, Manuela Födinger, Johannes Zuber, Vienna Covid-19 Detection Initiative (VCDI), Franz Allerberger, Andrea Pauli, Julius Brennecke

**Affiliations:** Research Institute of Molecular Pathology (IMP), Vienna BioCenter (VBC), Campus-Vienna-Biocenter 1, 1030 Vienna, Austria; Institute of Molecular Biotechnology of the Austrian Academy of Sciences (IMBA), Vienna BioCenter (VBC), Dr. Bohr-Gasse 3, 1030 Vienna, Austria; Vienna BioCenter PhD Program, Doctoral School of the University at Vienna and Medical University of Vienna, Vienna, Austria; Institute for Medical Microbiology and Hygiene, Austrian Agency for Health and Food Safety, Währingerstr. 25A, 1090 Vienna, Austria; European Centre for Disease Prevention and Control (ECDC), Stockholm, Sweden; Institute of Laboratory Diagnostics, Klinik Favoriten, 1100 Vienna, Austria; Sigmund Freud Private University, 1020 Vienna, Austria; Medical University of Vienna, Vienna BioCenter (VBC), 1030 Vienna, Austria; Gregor Mendel Institute (GMI), Austrian Academy of Sciences, Vienna Biocenter (VBC), Dr. Bohr-Gasse 3, 1030 Vienna, Austria; Max Perutz Labs, Medical University of Vienna, Vienna Biocenter (VBC), Dr. Bohr-Gasse 9/3, 1030 Vienna, Austria; Vienna Biocenter Core Facilities GmbH (VBCF), Dr. Bohr-Gasse 3, 1030 Vienna, Austria; Centre for Microbiology and Environmental Systems Science, University of Vienna, Althanstrasse 14, 1090 Vienna, Austria; Joint Microbiome Facility of the University of Vienna and Medical University of Vienna, Althanstrasse 14, 1090 Vienna, Austria; Institute of Biochemistry, University of Natural Resources and Life Sciences (BOKU), Muthgasse 18, 1190 Vienna, Austria; Department of Microbiology, Immunobiology, and Genetics, Max Perutz Labs, University of Vienna, Vienna Biocenter (VBC), Dr. Bohr-Gasse 9, 1030 Vienna, Austria

**Keywords:** Covid-19 diagnostics, RT-LAMP, RT-PCR, Coronavirus, SARS-CoV-2

## Abstract

Viral pandemics, such as Covid-19, pose serious threats to human societies. To control the spread of highly contagious viruses such as SARS-CoV-2, effective test-trace-isolate strategies require population-wide, systematic testing. Currently, RT-qPCR on extracted RNA is the only broadly accepted test for SARS-CoV-2 diagnostics, which bears the risk of supply chain bottlenecks, often exaggerated by dependencies on proprietary reagents. Here, we directly compare the performance of gold standard diagnostic RT-qPCR on extracted RNA to direct input RT-PCR, RT-LAMP and bead-LAMP on 384 primary patient samples collected from individuals with suspected Covid-19 infection. With a simple five minute crude sample inactivation step and one hour of total reaction time, we achieve assay sensitivities of 98% (direct RT-PCR), 93% (bead-LAMP) and 82% (RT-LAMP) for clinically relevant samples (diagnostic RT-qPCR Ct <35) and a specificity of >98%. For direct RT-PCR, our data further demonstrate a perfect agreement between real-time and end-point measurements, which allow a simple binary classification similar to the powerful visual readout of colorimetric LAMP assays. Our study provides highly sensitive and specific, easy to implement, rapid and cost-effective alternatives to diagnostic RT-qPCR tests.

## RESULTS & DISCUSSION

We obtained 384 primary patient samples to benchmark, in parallel, three laboratory-developed SARS-CoV-2 tests [1] against a diagnostic RT-qPCR assay, which uses extracted RNA as input [2] (Figure 1A). 271 samples were naso-oropharyngeal swabs in viral transport medium (VTM) or 0.9% NaCl solution (saline), and 113 were gargle samples in saline or Hank’s Balanced Salt Solution (HBSS). For all samples, we quantitatively determined SARS-CoV-2 levels via the gold-standard RT-qPCR (E-gene) on extracted RNA in the diagnostic laboratory of the Austrian Agency for Health and Food Safety (AGES), the Austrian national public health agency. This confirmed SARS-CoV-2 virus in 161/384 samples (102 swabs and 59 gargle specimens) with viral titres ranging between 0.2 and 100 million copies per microliter primary sample (Supplementary Figure S1A). Each of the three rapid assays described below direct RT-PCR, RT-LAMP, bead-LAMP bypasses the time-consuming and costly RNA extraction step. Instead, we used a five-minute inactivation procedure of the primary sample in QuickExtract (Lucigen) buffer, which effectively lyses virions and permanently inactivates RNases, thereby generating a non-infectious sample with stable viral RNA [3]. This method is compatible with naso-oropharyngeal swabs, sputum, saliva and gargle [1,4].

**Figure 1:**
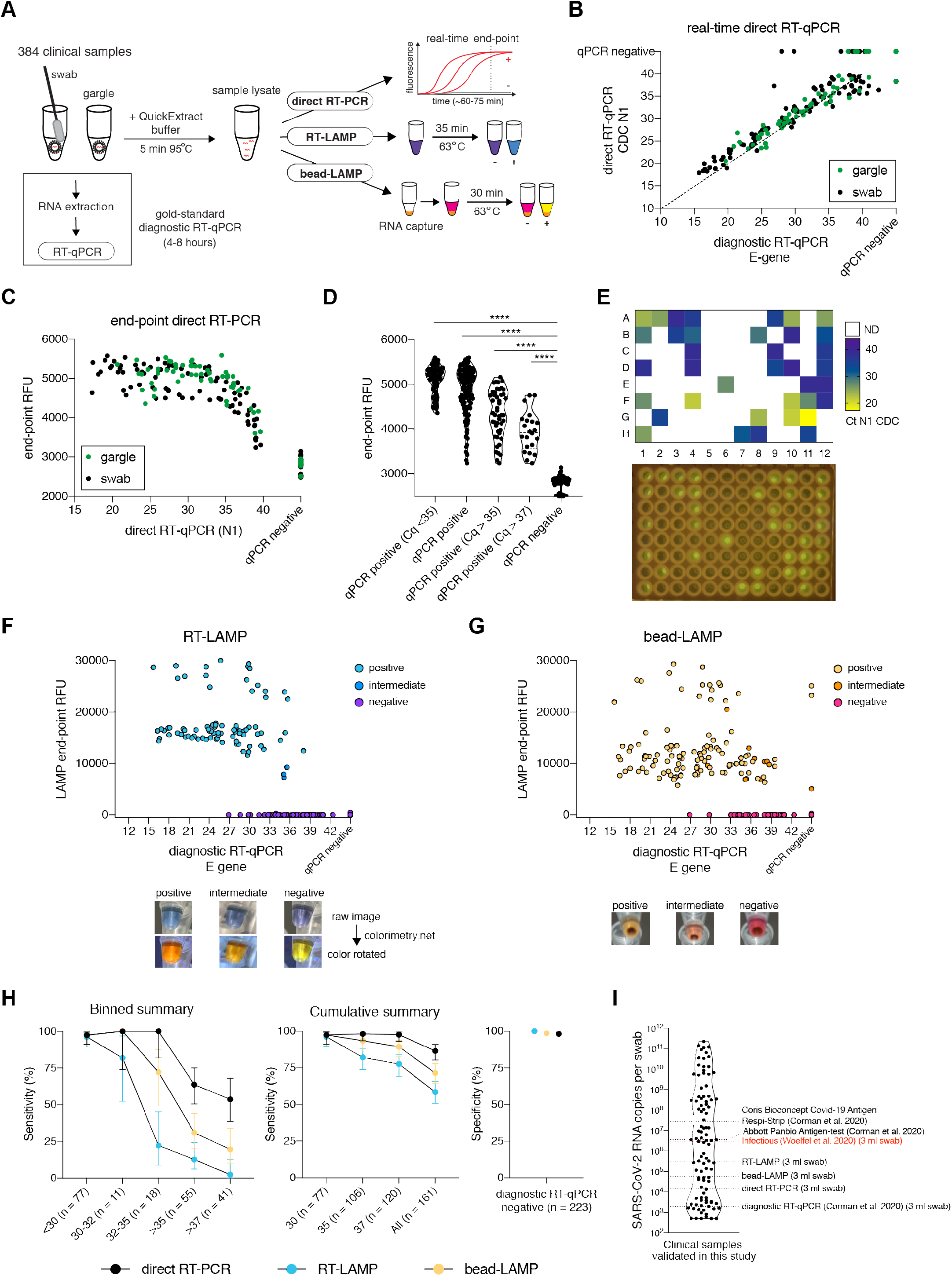
Head-to-head comparison of direct-input RT-PCR and RT-LAMP assays against diagnostic RT-qPCR. **A**. Schematic illustrating the workflow for the head-to-head comparison of three rapid, laboratory-developed tests (direct RT-PCR, RT-LAMP, bead-LAMP) against diagnostic RT-qPCR for rapid SARS-CoV-2 diagnostics. A five-minute heat-inactivation step in QuickExtract buffer circumvents the RNA extraction of the gold-standard diagnostic RT-qPCR protocol. **B**. Comparison of real-time direct RT-qPCR to diagnostic RT-qPCR. Each dot represents the Ct (cycle threshold) value of an individual sample measured by the two techniques. The color indicates the sample type (naso/oropharyngeal swab or gargle). The dashed line indicates a 100% theoretical agreement. Spearman’s correlation coefficient (Rho) was calculated for the correlation analysis between diagnostic RT-qPCR and direct RT-qPCR. **C**. Comparison of real-time direct RT-qPCR and end-point direct RT-PCR. Each dot represents the Ct value of an individual sample and its corresponding end-point relative fluorescence units (RFU) measured from the same reaction. Sample type is indicated by the color of the data-point (naso/ oropharyngeal swab or gargle). **D**. Data shown in C) split up into groups by diagnostic RT-qPCR Ct values. Mann-Whitney test was used to compare end-point RFUs within individual groups to the values measured for RT-qPCR negative samples (**** < 0.0001 significance). **E**. Heatmap showing measured direct RT-qPCR Ct values for a 96-well sample plate (top) and the corresponding end-point fluorescence smartphone image taken on a standard LED blue-light transilluminator (bottom). **F**. Comparison of RT-LAMP to diagnostic RT-qPCR. Each dot represents the Ct value of an individual sample and its corresponding end-point RFU values measured by RT-LAMP reactions containing HNB and the fluorescent SYTO-9 dye. The color of each data point indicates the final reaction color obtained by RT-LAMP. Representative examples of final reaction colors (raw images) and the corresponding color-rotated images (colorimetry.net), which facilitate scoring of the color changes, are shown underneath. **G**. Same as shown in F) but for Kingfisher bead-LAMP using 60 µl of lysate as input. **H**. Determined assay sensitivity and specificity of direct RT-PCR (black), RT-LAMP (blue) and bead-LAMP (yellow) in comparison to diagnostic RT-qPCR performed on the same 384 samples. The binned (left) and cumulative (middle) sensitivity graphs show the mean detection rate with error bars representing the 95% confidence interval (Wilson/Brown binomial confidence interval). The assay specificity graph (right) shows the percentages of negative agreement between each of the alternative tests with the diagnostic RT-qPCR. **I**. Overview of different SARS-CoV-2 detection assays and their respective limits of detection. The SARS-CoV-2 RNA concentration for each clinical naso/oro-pharyngeal swab sample was calculated from diagnostic RT-qPCR Ct values and plotted (log10). Dotted lines indicate the limit of detection (95% confidence interval) for various assays measured in this study (RT-LAMP, bead-LAMP, direct RT-PCR) or elsewhere (Abbott Panbio Antigen-test and Coris Bioconcept Covid-19 Antigen Respi-Strip [13], clinical RT-qPCR [2]). A viral titre at or above the indicated “infectious” line (red) has previously been determined in a virological assessment study of hospital patients [11].

### Direct RT-PCR

The direct RT-qPCR method is analogous to diagnostic RT-qPCR assays but circumvents the dedicated RNA extraction step and uses inexpensive, readily available reagents [3,5]. We performed direct RT-qPCR using Luna One-Step RT-qPCR reagents (New England Biolabs) and the CDC N1 SARS-CoV-2 reference primer/probe set (IDT) [1]. The obtained Ct-values correlated strongly with the diagnostic RT-qPCR values (Spearman correlation Rho = 0.93), demonstrating that omitting the RNA extraction step does not distort quantitative measurements and that RT-PCR-inhibiting substances are not prevalent in naso-oropharyngeal swab and gargle samples (Figure 1B). On average, Ct values of the direct RT-PCR assay were ∼2 cycles higher compared to the diagnostic Ct-values. This indicated minor losses in sensitivity (Figure 1B), which we attribute to the lower input amount per reaction in direct RT-PCR (no RNA extraction and enrichment compared to the diagnostic RT-qPCR). With the exception of two outliers (Ct 28 and Ct 29), all samples with a diagnostic RT-qPCR Ct value below or equal to 35 (more than ∼30 viral copies per reaction; n = 104 samples) as well as 35 out of 55 samples with clinical Ct values above 35 (63.3%) were positive by direct RT-qPCR (Figure 1B, H). Thus, specimens yielding false negative results by the direct RT-qPCR method had very low viral copy numbers (Figure 1B, H). Among the samples yielding false negative results, gargle and swab specimens showed no statistically significant difference in sample Ct (P = 0.89, Mann-Whitney test) or Ct value distribution (P_gargle_ = 0.27, P_swab_ = 0.17, Kolmogorov–Smirnov test) (Supplementary Figure S1B). Direct RT-qPCR detected SARS-CoV-2 in four specimens defined as ‘negative’ by the diagnostic RT-qPCR assay (Figure 1B). The estimated viral loads of these samples were at the limit of detection by diagnostic RT-qPCR (Ct above 38, less than ∼5 copies per reaction). Given the otherwise high specificity of the direct RT-PCR method (98.2% negative predictive agreement for 223 samples; Figure 1H), we suspect minor cross-contamination from positive samples as the likely cause (e.g. three out of the four samples were adjacent to SARS-CoV-2 positive sample wells).

When using real-time qPCR instruments, the direct RT-qPCR assay is quantitative in nature (Figure 1B). For a simple virus-positive or negative classification, however, an end-point measurement using a standard laboratory PCR machine and a simple illumination device would be sufficient. To test this, we performed end-point fluorescence measurements of RT-qPCR reactions that were also monitored in real-time. This revealed a clear quantitative difference in relative fluorescence units (RFU) between RT-qPCR-positive and -negative samples (P < 0.0001, Mann-Whitney test) (Figure 1C, D). Primary samples with diagnostic Ct values below or equal to 35 (more than ∼30 viral copies per reaction) displayed the strongest signal intensity (RFU_max_ = 5590), and even weakly positive samples with Ct values above 37 (less than ∼5 copies per reaction) displayed RFU values greater than the maximum measured for any of the 240 negative samples (Figure 1C, D). Endpoint RT-PCR therefore allows a robust, automated yes/no classification based on co-measured positive and negative control reactions. The strong fluorescent signal is visible in PCR plates or strips when using a standard blue-light transilluminator or a simple blue light LED lamp (Figure 1E, Supplementary Figure S1C), and images can be captured with a standard smartphone camera. We observed 100% concordance between machine-read (RT-qPCR cycler or plate reader) and visible fluorescence (Figure 1E, Supplementary Figure S1C). For an automated, rapid classification of the assay, fluorescent images can be converted into signal intensities using the FIJI6 plugin ReadPlate (Supplementary Figure S1D).

### RT-LAMP

Isothermal, loop-mediated amplification after reverse-transcription (RT-LAMP) is a powerful nucleic acid amplification method [7] that requires minimal equipment (in its simplest form, a water bath at 62 - 65°C [1]; Figure 1A, Supplementary Figure S 2 A, B). RT-LAMP based DNA amplification is rapid (< 30 minutes) and, due to the large amount of generated DNA products, allows the use of a colorimetric and/or fluorescence readout [8]. Instead of the pH-dependent dye Phenol Red commonly used in RT-LAMP assays, we chose the metal-indicator HNB [9] as it is compatible with the QuickExtract buffer and all commonly used specimen types (like swabs, gargle samples, saliva, sputum) and media (VTM, saline, HBSS) [1,4]. To simultaneously detect fluorescence, we used the SYTO-9 dye (detailed LAMP protocol available at https://www.rtlamp.org/). From the 384 patient samples, RT-LAMP targeting the SARS-CoV-2 ORF1ab gene [10] detected 94 out of the 161 RT-qPCR positive samples (58%) while maintaining a 100% negative predictive agreement rate with no false positive results among the 223 specimens that were determine ‘negative’ by diagnostic RT-qPCR (Figure 1F, H). Concordance between colorimetric results (sky-blue = positive, purple = negative) and fluorescence intensity (LAMP end-point RFU) was 100%, demonstrating the reliability in detecting positive specimens based on colorimetric signal alone (Figure 1F, Supplementary Figure S3). The assay sensitivity for samples with a diagnostic RT-qPCR Ct value below or equal to 30 was 96.1% (74/77), 44.8% for Ct values between 30 to 35 (13/29), and 12.7% for weakly positive samples with Ct values above or equal to 35 (7/55) (Figure 1F, H). The mean time-to-threshold (time until fluorescent signal reached a threshold value) was ∼15 minutes (range from 9.9 to 23.3 minutes) (Supplementary Figure S2C). Reactions displaying an intermediate colour had time to threshold values of more than 20 minutes (Figure 1F, Supplementary Figure S2C), indicating that for lower viral loads (RT-qPCR Ct values above 33) fluorescence is the more robust readout. Thus, RT-LAMP is a very fast, simple and versatile assay that has a sensitivity sufficient to identify SARS-CoV-2 in samples from infectious individuals [11] (Figure 1I).

### Bead-LAMP

To increase the sensitivity of RT-LAMP, we developed bead-enriched RT-LAMP (in short, bead-LAMP), which uses a 15-minute, magnetic bead based enrichment step to capture RNA from up to 100 µl inactivated patient samples (Figure 1A, Supplementary Figure S2B) [1]. RNA elution in bead-LAMP is done directly with the RT-LAMP reaction mix, increasing speed and allowing for maximum RNA enrichment. As bead-enrichment removes sample buffer and matrix components, it is compatible with Phenol Red-based colorimetric detection (pink = negative; yellow = positive) (Figure 1G, Supplementary Figure S2B, Supplementary Figure S3; for detailed bead-LAMP protocols visit https://www.rtlamp.org/). Bead-LAMP can be performed manually as well as on the commonly used, fully automated Kingfisher Flex purification system, which reduces the amount of valuable reaction consumables such as pipette tips. Bead-LAMP detected 115 out of 161 RT-qPCR positive samples (71.4%) (Figure 1G). As expected, bead-LAMP improved assay sensitivity, detecting 83% (30/36) of samples with diagnostic Ct values between 30 and 35, 31% for Ct values above 35 (17/55) and even 19.5% (8/41) of samples with Ct values above 37 (Figure 1G, H). In comparison, direct RT-LAMP detected only 1 out of 41 samples in the Ct range above 37 (Figure 1F, H). Out of 223 specimens that yielded negative results by RT-qPCR, three specimens (1.3%) showed positive results when tested by bead-LAMP (Figure 1G). None of these three samples were positive in direct RT-qPCR or RT-LAMP, suggesting that they were in fact incorrect positive results of the bead-LAMP assay. We suspect that minor cross-contaminations during sample handling are the underlying cause of the false positive results. Compared to the manual bead-LAMP protocol, the automated KingFisher system improved the overall performance of bead-LAMP despite using lower sample input volume per sample (60 µl versus 100 µl). We measured a subset of samples (n = 288) in parallel using manual and automated bead-LAMP and observed higher detection rates in the fully automated KingFisher version (Supplementary Figure S2D, S3). As for RT-LAMP, no difference in sensitivity was observed for bead-LAMP between gargle and swab samples (Supplementary Figure S2E), demonstrating compatibility with diverse input samples. Taken together, bead-LAMP is an attractive solution for applications where increased assay sensitivity is required, such as for pooled testing [1].

Our comparative study highlights three laboratory-developed SARS-CoV-2 tests that are cheap (reagent costs ∼1 USD for RT-qPCR and RT-LAMP, ∼1.2 USD for bead-LAMP), rapid (45 - 80 min total test time), simple, and that circumvent supply chain bottlenecks. The three assays hold the potential to greatly expand the availability of affordable testing strategies anywhere in the world. Our data demonstrate that direct RT-PCR, RT-LAMP and bead-LAMP are fully compatible with crude sample lysates without measurable reaction inhibition. In line with previous studies [5,10,12], this strongly supports the validity of extraction-free nucleic acid diagnostics. All three assays are inherently scalable and have a sensitivity that exceeds the required threshold to detect infectious individuals, which only the best commercial rapid Antigen point-of-care tests achieve [13] (Figure 1I). All three assays are compatible with various input types such as swabs or gargle, the latter offering a simple, non-invasive way of self-collecting the specimen without the need of trained, specialized personell. Finally, all three of the rapid assays highlighted here take advantage of the dUTP/UDG system [14-16], which we used throughout this study in order to minimize technical false-positive results due to carry-over contamination with prior PCR or LAMP amplicons.

Depending on the available equipment and desired assay sensitivity, our nucleic acid amplification assays provide an effective alternative to diagnostic RT-qPCR testing, where dependencies on commercial testing platforms and reagent supply shortages could lead to unused testing capacities. Laboratories already performing RT-qPCR or regular PCRs could implement direct RT-qPCR or end-point RT-PCR, while our end-point RT-PCR and colorimetric RT-LAMP protocols provide powerful, simple testing methods to greatly increase the testing capacity particularly in settings where qPCR detection instruments are unavailable. For settings in which speed and simplicity matters, colorimetric RT-LAMP is the most powerful method. Furthermore, the sensitivity of RT-PCR for pooled tests, and the speed of RT-LAMP for de-pooling could be combined to massively scale up testing in cases of low viral prevalence in the population [17]. In this, our rapid and cost-effective methods could enable any country to establish population-wide testing strategies, which would be highly effective in limiting the spread of contagious viruses such as SARS-CoV-2 [18,19].

## Data Availability

All data is available within the submitted manuscript.

## ACKNOWLEDGMENTS

This work would not have been possible without the enthusiastic support of the IMBA and IMP research institutes, as well as the many volunteers and partners of the VCDI, who came together to help and collaborate under the exceptional circumstances of the Covid-19 pandemic. Special thanks go to H. Isemann for his administrative and overall support, as well as to the Covid-19 NGS team (R. Yelagandula, L. Cochella and Uli Elling) for discussions and experimental advice. We thank the Pauli and Brennecke groups for bearing with us and sharing lab space. We thank Nathan Tanner (NEB) for valuable discussions and sharing important information on LAMP technology. We are grateful to the Covid Testing Scaleup SLACK channel for openly sharing and exchanging information and the Covid-19 diagnostics team in Feng Zhang’s lab for discussions.

## AUTHOR CONTRIBUTIONS

MJK and MM designed, performed and analyzed all experiments. MJK, MM, JJR, JSchnabl and DH developed LAMP and direct-PCR assays. RH established the KingFisher protocol. FA, AI, PH and JSchaeffer coordinated patient samples and provided clinical RT-qPCR measurements. MF, RFP, JZ and MPSD provided and helped with patient samples for method development. JZ acquired project funding. The VCDI supported Covid-19 testing initiatives at the Vienna BioCenter. MJK, AP and JB conceived the project. AP and JB supervised the project. MJK, MM, AP and JB wrote the paper with input from all authors.

## MATERIALS AND METHODS

### Clinical sample collection

Patient samples (oro/nasopharyngeal swabs and gargle) were obtained as part of a clinical performance study approved by the local Ethics Committee of the City of Vienna (#EK 20-292-1120). Oro/Nasopharyngeal swabs were collected in 3 ml Viral Transport Medium (VTM) or 0.9% NaCl solution (saline). Gargle samples were collected from patients by letting individuals gargle for 1 minute with 5 ml of Hank’s Balanced Salt Solution (HBSS) or 0.9% saline solution. Informed consent was obtained from all patients.

### RNA extraction and clinical RT-qPCR

RNA was extracted from 200 µl of NP and OP swab supernatants using a commercial kit (BioExtract® SuperBall®, BioSellal, France) and the KingFisher™ Flex Purification System (Thermo Fisher Scientific, USA). Negative extraction controls (nuclease-free water) were prepared alongside clinical samples to monitor for potential cross-contamination. Detection of SARS-CoV-2 RNA was performed using a commercial primer/probe mix (LightMix® Modular SARS and Wuhan CoV E-gene; TIB Molbiol, Germany) and Super Script™ I I I Platinum® One-Step Quantitative RT-PCR System with ROX (Thermo Fisher Scientific, USA) on the ABI7500Fast system (Thermo Fisher Scientific, USA). Assays were performed as in duplicate as real-time RT-PCR reactions, also targeting β-actin mRNA as extraction control [20]. Nuclease-free water and a synthetic RNA control provided with the primer/probe mix were included as respective no-template-control (NTC) and positive control (PC). A CT-value of >40 was considered a negative result.

### Crude sample inactivation using QuickExtract DNA solution

120 µl of naso/oro-pharyngeal swab or gargle sample was mixed 1:1 with QuickExtract™ DNA Extraction Solution (Lucigen) and heat inactivated for 5 minutes at 95°C. Samples were then stored on ice until further use or frozen at −80°C.

### Direct RT-PCR

For detecting the viral N-gene via RT-qPCR, 1-step RT-qPCR was performed using the Luna Universal One-Step RT-qPCR Kit (NEB), 1.5 µl of reference primer/probe sets CDC-N1 (IDT 10006713), 0.4 µl of Antarctic Thermolabile UDG (NEB), and 2 µl of QuickExtract inactivated crude sample lysate per 20 µl reaction. Reactions were run at 55°C for 10 minutes, 95°C for 1 minute, followed by 45 cycles of 95°C for 10 seconds and 55°C for 30 seconds in a Bio Rad CFX q PCR cycler. End-point measurements were taken on the BioRad CFX qPCR cycler after 45 cycles of amplification and plotted without baseline subtraction. Alternatively, PCR plates were placed on a LED Blue-light Transilluminator (Safe Imager 2.0, Thermofisher) and photographed using a regular smartphone camera (iPhone XS). The Fiji plugin ReadPlate was used to convert the smartphone image into numerical values according to the plugin’s manual instructions.

### RT-LAMP

For detailed, up-to-date protocols on the RT-LAMP assay visit https://www.rtlamp.org/get-started/rt-lamp-commercial/. In brief, RT-LAMP reactions were set up by mixing 10 µl of the 2X NEB WarmStart® RT-LAMP Master Mix (1x final, 2x stock) with 2 µl of 10x ORF1a-HMS primer solution [10] (1x final equals 1.6 µM of FIP, 1.6 µM of BIP, 0.4 µM of LB, 0.4 µM of LF, 0.2 µM of F3, 0.3 µM of B3), 0.4 µl of 100 µM SYTO-9 dye (Thermofisher, made from 5 mM Stock in DMSO), 0.4 µl of NEB Antarctic thermolabile UDG, 0.14 µl of 100 mM dUTP solution (NEB, 0.7 mM final dUTP added), 0.14 µl of 100 mM MgSO4 (NEB, 0.7 mM added), 0.8 µl of 3 mM hydroxynapthol blue (120 µM final, stock solution made from HNB powder (Hach) in nuclease-free water and filtered through 0.22 µm filter), 2 µl of QuickExtract inactivated crude sample lysate and nuclease-free water to a final volume of 20 µl. Reactions were run at 63°C for 35 cycles (1 minute cycle length) in a BioRad CFX Connect qPCR cycler with SYBR readings after every cycle. Colorimetric signal was read out after reaction completion using a simple smartphone camera. Reactions with sky blue color were scored as positive, purple color as negative and everything between as intermediate. A separate classification was done to compare results using a color conversion tool (colorimetry.net) which rotates the color space by 180° to invert reaction colors into orange (positive) or yellow (negative).

### Bead-LAMP

For detailed, up-to-date protocols on the bead-LAMP assay visit https://www.rtlamp.org/get-started/bead-lamp-commercial/. In brief, for bead enrichment, 60 µl or 100 µl of QuickExtract inactivated crude lysate was mixed with 0.6x of magnetic beads (1:5 dilution of Agencourt RNAClean XP in 2.5 M NaCl, 10 mM Tris-HCl pH 8.0, 20% (w/v) PEG 8000, 0.05% Tween 20, 5 mM NaN3) and either manually or automatically processed. For manual bead-LAMP, bead-sample mixtures were incubated for 5 minutes at room temperature followed by bead capture on a magnetic rack (Alpaqua 96S Super Magnet) for 5 minutes. The liquid was removed and beads were washed twice with 85% ethanol for 30 seconds. The beads were air dried for 5 minutes and then eluted directly in 20 µl colorimetric LAMP reaction mix containing 10 µl of 2x NEB WarmStart® Colorimetric LAMP Master Mix with UDG (1x final), 2 µl of 10x ORF1a-HMS [10] and 2 µl of NEB E1 primer solution [21] (1x final equals 1.6 µM of FIP, 1.6 µM of BIP, 0.4 µM of LB, 0.4 µM of LF, 0.2 µM of F3, 0.3 µM of B3), 0.4 µl of 2M GuHCl (40 mM final, 2M Stock made from pH 8.5 buffered solution in water (Sigma G7294)), 0.4 µl of 100 µM SYTO-9 dye (ThermoFisher, made from 5 mM Stock in DMSO) and nuclease-free water to a final volume of 20 µl. For automated bead-LAMP, we established a custom protocol for the KingFisher™ Flex Purification System: The protocol was designed for a 96-well PCR magnetic head and maximum 100 µl of total sample volume. After mixing 60 µl of crude lysate with 40 µl of magnetic beads (1:5 dilution of Agencourt RNAClean XP in 2.5 M NaCl, 10 mM Tris-HCl pH 8.0, 20% (w/v) PEG 8000, 0.05% Tween 20, 5 mM NaN3), the sample plate was placed into the KingFisher system. The protocol was initiated by mixing for 5 minutes at medium speed. Next, beads were collected through 5 iterations of 30 seconds each. Then beads were washed for 15 seconds in a wash plate containing 85% EtOH. For this, a bead-release step of 15 seconds was done in the beginning of the wash step, followed by collecting the beads three times for 5 seconds. Beads were then air-dried for 3 minute and released into the bead-LAMP reaction mixture for 1 minute at fast speed. The protocol is available upon request.

Both manual and automated KingFisher bead-LAMP reactions were run at 63°C for 25 cycles (1 minute cycle length) in a BioRad CFX Connect qPCR cycler with SYBR readings after every cycle. Colorimetric signal was read out after reaction completion using a simple smartphone camera. Reactions with yellow color were scored as positive, pink color as negative and everything between as intermediate.

### LAMP primer sequences

#### ORF1a-HMS

ORF1a-HMS_F3

CGGTGGACAAATTGTCAC

ORF1a-HMS_B3:

CTTCTCTGGATTTAACACACTT

ORF1a-HMS_LF:

TTACAAGCTTAAAGAATGTCTGAACACT

ORF1a-HMS_LB:

TTGAATTTAGGTGAAACATTTGTCACG

ORF1a-HMS_FIP:

TCAGCACACAAAGCCAAAAATTTATCTGTGCAAAGGAAATTAAGGAG

ORF1a-HMS_BIP:

TATTGGTGGAGCTAAACTTAAAGCCCTGTACAATCCCTTTGAGTG

#### NEB-E1

NEB-E1_F3:

TGAGTACGAACTTATGTACTCAT

NEB-E1_B3:

TTCAGATTTTTAACACGAGAGT

NEB-E1_FIP:

ACCACGAAAGCAAGAAAAAGAAGTTCGTTTCGGAAGAGACAG

NEB-E1_BIP:

TTGCTAGTTACACTAGCCATCCTTAGGTTTTACAAGACTCACGT

NEB-E1_LB:

GCGCTTCGATTGTGTGCGT

NEB-E1_LF:

CGCTATTAACTATTAACG

## SUPPLEMENTARY FIGURES

**Figure S1:**
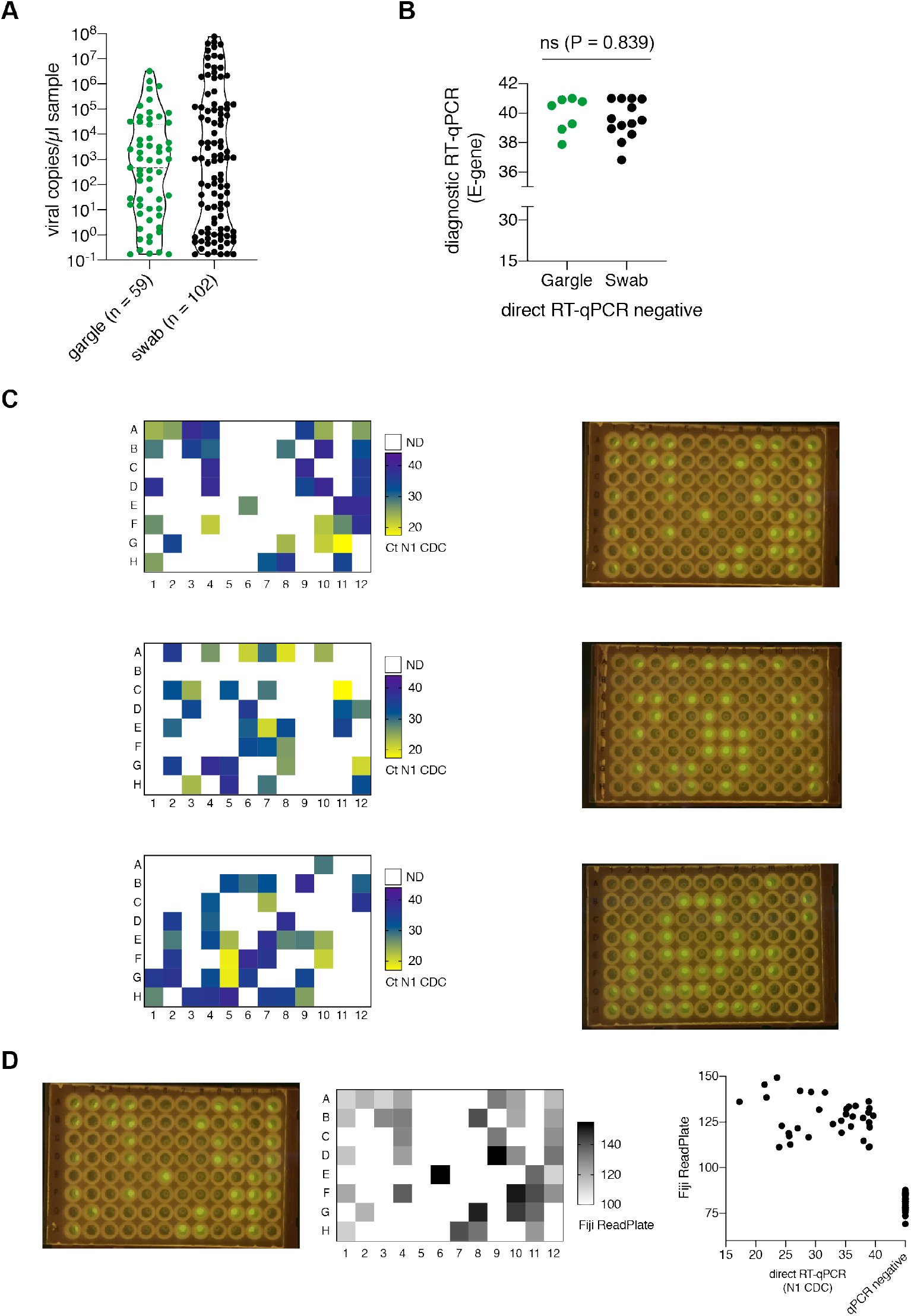
Real-time and end-point fluorescence direct RT-PCR. **A**. Violin plot showing estimated SARS-CoV-2 RNA copies per clinical sample (naso/oropharyngeal swab or gargle). A standard curve was used to convert Ct values into viral copies per reaction and projected to the total sample volume (3 ml for naso/oropharyngeal swab, 5 ml for gargle). **B**. Comparison of diagnostic RT-qPCR Ct values that were not detected by direct RT-qPCR by sample type (gargle versus naso/oropharyngeal swab). No significant (ns) difference was observed between the two groups (P = 0.839, Mann-Whitney test). **C**. Heatmap showing measured direct RT-qPCR Ct values for three different sample plates (left) and the corresponding end-point fluorescence smartphone images taken on a standard LED blue-light transilluminator (right). **D**. Semi-automated classification of end-point fluorescence smartphone images by digitalisation of the raw fluorescence images. The smartphone image (left) is analysed using the Fiji ReadPlate plugin, which converts fluorescence intensities into numerical values. The numerical values are shown as heatmap in plate-format (middle) or scatterplot with the corresponding direct RT-qPCR Ct values (right).

**Figure S2:**
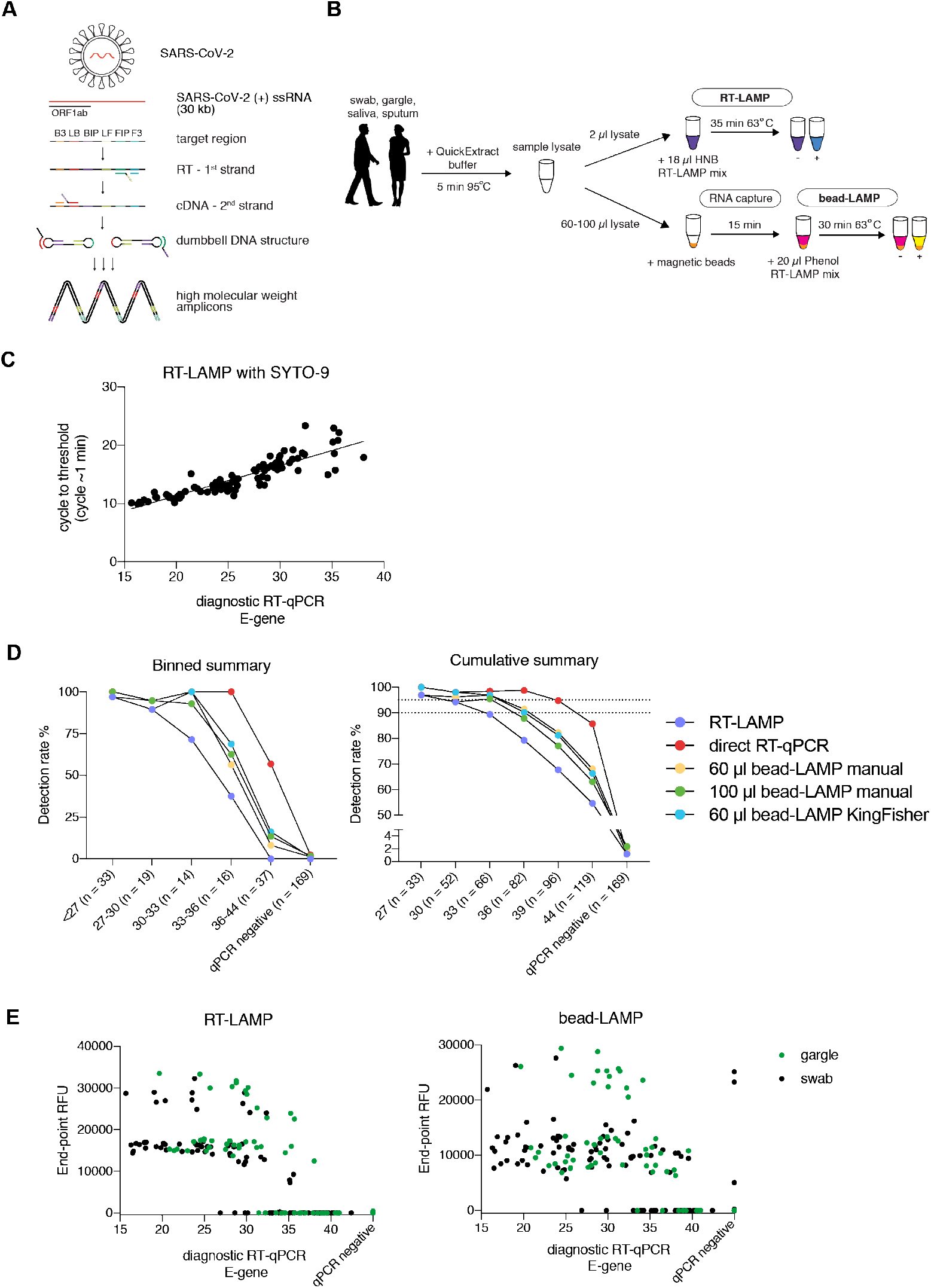
RT-LAMP and bead-LAMP. **A**. Schematic illustrating the reaction principle of reverse transcription loop-mediated amplification (RT-LAMP) of SARS-CoV-2 RNA and the RNA region targeted in this study (Orf1ab). The target region is recognized by a defined set of primers (B3, LB, BIP, LF, FIP, F3). The RNA template (red) is reverse transcribed and displaced after first-strand synthesis; the outer primer binding sites are added in the subsequent amplification step. The resulting dumbbell DNA structure acts as template for further rounds of amplification, ultimately leading to high molecular weight amplicons. **B**. Schematic illustrating the workflow for the clinical assessment of RT-LAMP using HNB colorimetric read-out (top) or bead-LAMP using Phenol-red colorimetric read-out (bottom). Bead-LAMP includes an upstream step, in which RNA is captured from crude lysate using magnetic beads, followed by elution by the RT-LAMP reaction mixture [1]. C. Scatterplot comparing RT-LAMP reaction times to direct RT-qPCR Ct values for each sample. The RT-LAMP reaction time was obtained by real-time fluorescence RT-LAMP, using SYTO-9 as fluorescent dye, and corresponds to the time at which the fluorescence RT-LAMP signal crosses the signal threshold. **D**. Determined binned (left) and cumulative (right) assay sensitivity of RT-LAMP (blue), bead-LAMP (yellow) and direct RT-PCR on 288 individual clinical samples. For bead-LAMP, both manual and automated (KingFisher) protocol performances using different amounts of sample input are shown. **E**. Scatterplots comparing real-time diagnostic RT-qPCR Ct values (x-axis) to RT-LAMP relative fluorescence units (y-axis). The color indicates the sample type (naso/oropharyngeal swabs or gargle). RT-LAMP is shown on the left, bead-LAMP is shown on the right.

**Figure S3:**
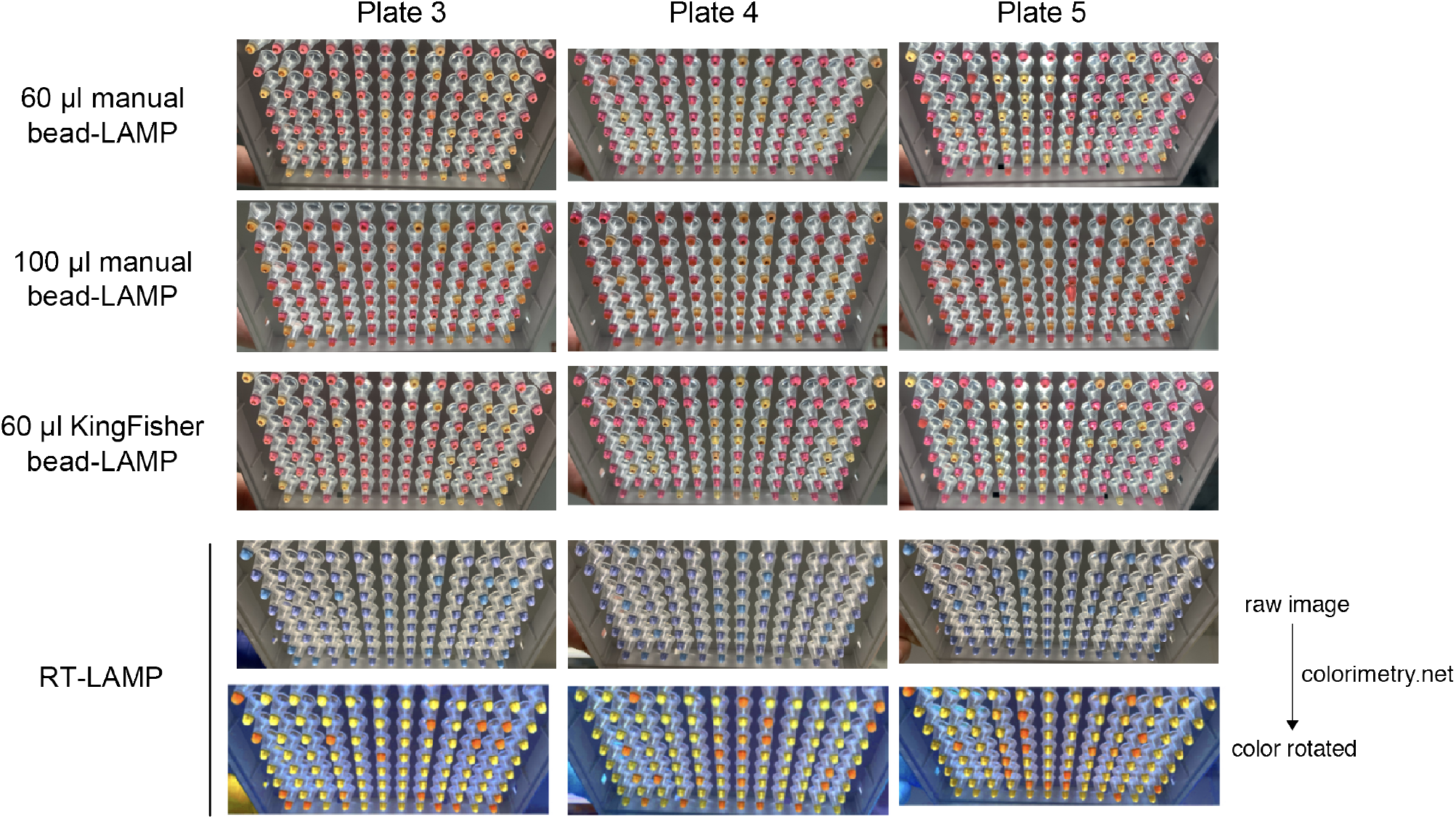
Colorimetric read-out of RT-LAMP and bead-LAMP. Shown are images from bead-LAMP and RT-LAMP after reaction completion for three independent sample plates (columns). For bead-LAMP, plate images for different lysate amounts as well as the performed protocol (manual versus automated KingFisher) are shown. To facilitate sample classification of colorimetric RT-LAMP reactions using the HNB dye, color converted images (www.colorimetry.net) are depicted underneath the original images obtained for those reactions.

